# NLRP3 inflammasome activation and altered mitophagy are key pathways in inclusion body myositis

**DOI:** 10.1101/2024.06.15.24308845

**Authors:** Elie Naddaf, Thi Kim Oanh Nguyen, Jens O. Watzlawik, Huanyao Gao, Xu Hou, Fabienne C. Fiesel, Jay Mandrekar, Eileen Kokesh, William S. Harmsen, Ian R. Lanza, Wolfdieter Springer, Eugenia Trushina

**Author notes:** **Corresponding Author:** Elie Naddaf, MD, Mayo Clinic Department of Neurology 200 First Street SW, Rochester, MN 55905.

## Abstract

**Background:** Inclusion body myositis (IBM) is the most prevalent muscle disease in adults for which no current treatment exists. The pathogenesis of IBM remains poorly defined. Inflammation and mitochondrial dysfunction are the most common histopathological findings. In this study, we aimed to explore the interplay between inflammation and mitochondrial dysfunction in IBM patients, highlighting sex differences.

**Methods:** We included 38 IBM patients and 22 age- and sex-matched controls without myopathy. Bulk RNA sequencing, Meso Scale Discovery ELISA, western blotting, histochemistry and immunohistochemistry were performed on frozen muscle samples from the study participants.

**Results:** We demonstrated activation of the NLRP3 inflammasome in IBM muscle samples, with the NLRP3 inflammasome pathway being the most upregulated. On muscle histopathology, there is increased NRLP3 immunoreactivity in both inflammatory cells and muscle fibers. Mitophagy is critical for removing damaged mitochondria and preventing the formation of a vicious cycle of mitochondrial dysfunction—NLRP3 activation. In the IBM muscle samples, we showed altered mitophagy, most significantly in males, with elevated levels of p-S65-Ubiquitin, a mitophagy marker. Furthermore, p-S65-Ubiquitin aggregates accumulated in muscle fibers that were mostly type 2 and devoid of cytochrome-c-oxidase reactivity. Type 2 muscle fibers are known to be more prone to mitochondrial dysfunction. *NLRP3 RNA* levels correlated with p-S65-Ubiquitin levels in both sexes but with loss of in muscle strength only in males. Finally, we identified sex-specific molecular pathways in IBM, with females having activation of pathways that could offset some of the pathomechanisms of IBM.

**Conclusions:** NLRP3 inflammasome is activated in IBM, along with altered mitophagy particularly in males, which is of potential therapeutic significance. These findings suggest sex-specific mechanisms in IBM that warrant further investigation.

## Introduction

Inclusion body myositis (IBM) is the most common muscle disease in aduls, for which there is no treatment.^1, 2^ It is characterized by relentlessly progressive muscle weakness, with decreased longevity and high morbidity as nearly all patients become wheelchair dependent as the disease progresses.^3, 4^ The pathomechanisms of IBM remain poorly understood posing major challenge for drug development. It is classified as an idiopathic inflammatory myopathy (IIM) mainly based on the associated histopathological findings, characterized by inflammatory infiltration of the endomysium, with clonally-restricted, highly differentiated, cytotoxic T cells.^5–7^ However, unlike any other IIM, IBM is a disease of aging that affects males twice as commonly as females, and does not respond to immunosuppressants or immunomodulatory drugs.^4, 8–10^ In addition to the inflammatory reaction, IBM is characterized by the accumulation of autophagic vacuoles and protein deposition such as amyloid-β peptides, phosphorylated tau protein, p62 and TDP43.^11, 12^ As these features are similar to other diseases of aging such as Alzheimer’s disease (AD) and Parkinson’s disease (PD), IBM is also considered a degenerative disease by some.^13^ Furthermore, the relationships among these various pathways remain poorly understood.

Histopathological evidence of mitochondrial dysfunction, as witnessed by the increased number of cytochrome c oxidase (CCO)-negative fibers, is observed in almost every IBM muscle biopsy. ^14–16^ In addition, multiple mitochondrial DNA deletions occur in IBM muscles, with greater mutation load in CCO-negative fibers. ^17, 18^ Mitochondrial dysfunction followed by endomysial inflammation are the two most commonly encountered histopathological features in IBM.^14, 15^ However, a role for mitochondrial autophagy (mitophagy) in IBM has not been established thus far. The best characterized stress-induced mitophagy pathway is jointly directed by the ubiquitin (Ub) kinase PINK1 and the Ub E3 ligase PRKN (parkin).^19^ In damaged mitochondria, PINK1 builds up on the outer mitochondrial membrane (OMM), activating the PINK1-PRKN enzymatic pair.^20^ PINK1 and PRKN then selectively decorate damaged mitochondria with phosphorylated (p-S65-) ubiquitin (Ub) chains that facilitate their degradation by the autophagy-lysosomal system.^21^ As such, the levels of p-S65-Ub are highly dynamic; they rise with the accumulation of p-S65-Ub up on damaged mitochondria, but then rapidly decline upon effective elimination of the organelles.

With the conundrum of involved pathways in IBM, systems biology approaches such as bulk transcriptomics analysis, are best suited as exploratory methods to study disease mechanisms. The results from previous transcriptomics studies performed on muscle samples from IBM patients were dominated by inflammation-related genes, highlighting pathways related to T-cell differentiation and cytotoxicity, interferon-ɣ signature, and immunoglobulin secretion.^7, 22–28^ The strong expression of inflammation-related genes posed a challenge to the exploration of noninflammatory pathways, which are essential for understanding IBM pathogenesis. Most importantly, sex differences in the transcriptome have never been addressed, despite the disease being more common and relatively more severe in males.^4, 8, 10, 29–31^ Hence, this study aimed to explore the interplay between mitochondrial dysfunction and inflammation in muscle tissue from male and female IBM patients, highlighting sex-specific differences.

## Methods

### Patient selection

A chart review of our electronic medical records was performed to identify IBM patients and controls. IBM diagnosis was established by fulfilling any category of the 2011 ENMC diagnostic criteria.^32^ Controls were identified from the Muscle laboratory database searchable by diagnosis. Patients were included if they had documented normal strength, normal creatine kinase levels, no fibrillation potentials on electromyography, and no myopathy or inflammation on muscle biopsy. Controls underwent a muscle biopsy to rule out a muscle disorder and the workup was unremarkable. The IBM patients and controls were age and sex matched at a group level. Disease duration was defined as the time from symptom onset until biopsy. The manual muscle testing (MMT) score is a summated motor exam of bilateral shoulder abduction, elbow flexion, elbow extension, finger flexion, hip flexion, knee extension and ankle dorsiflexion. Each muscle is graded from 0 (normal) to 4 (complete paralysis) and the total score ranges from 0 (normal strength) to 56 (complete paralysis) as previously described.^33, 34^ MMT was calculated at time of the biopsy. All muscle biopsies were originally obtained for clinical purposes per our clinical practice standards, and residuals were used for this project. After overnight fasting, an open biopsy was performed the following day in the operating room under conscious sedation. The harvested muscle tissue was frozen in isopentane cooled in liquid nitrogen and stored at −80°C according to our Muscle laboratory protocols.

### RNA extraction and sequencing

Approximately 20-30 mg of fresh frozen muscle tissue per sample were sent to GENEWIZ, LLC./Azenta US, Inc. (South Plainfield, NJ, USA) for RNA extraction and sequencing. Total RNA was extracted using Qiagen RNeasy Plus Universal kit following manufacturer’s instructions (Qiagen, Hilden, Germany). Quantification was performed using Qubit 2.0 fluorometer (Thermo Fisher Scientific, Waltham, MA, USA) and RNA integrity was checked using TapeStation (Agilent Technologies, Palo Alto, CA, USA). All included samples had a RIN>6. The RNA sequencing library was prepared using the NEBNext Ultra II RNA Library Prep Kit for Illumina using manufacturer’s instructions (New England Biolabs, Ipswich, MA, USA). The sequencing library was validated on the Agilent TapeStation (Agilent Technologies, Palo Alto, CA, USA), and quantified by using Qubit 2.0 fluorometer (Thermo Fisher Scientific, Waltham, MA, USA) as well as by quantitative PCR (KAPA Biosystems, Wilmington, MA, USA). The sequencing libraries were multiplexed and clustered across four flow cell lanes. After clustering, the flow cells were loaded onto the Illumina HiSeq instrument according to manufacturer’s instructions. The samples were sequenced using a 2×150 bp paired-end (PE) configuration. Image analysis and base calling were conducted by the HiSeq Control Software (HCS). The raw sequence data (.bcl files) generated from Illumina HiSeq were converted into fastq files and demultiplexed using Illumina bcl2fastq 2.20 software. One mismatch was allowed for index sequence identification.

### Transcriptomic data analysis

RNA sequencing data analysis was performed using Basepair software (https://www.basepairtech.com/) with a pipeline that included the following steps. The raw reads were trimmed using fastp to remove low-quality bases from the reads (quality < 10) and adapter sequences. Trimmed reads with a minimum length of 15 bp were aligned to the ENSEMBL GRCh38 genome assembly using STAR with default parameters.^35^ Alignments were sorted and indexed using samtools and normalized bigwig files were created for visualization using deepTools with a bin size of 5 bp. Read counts were collected at gene annotations using feature counts and data were visualized using the IGV browser. Differential expression analyses were performed using DESeq2. Pairwise comparisons were performed using the Wald test. The minimum gene expression cutoff was set at 10. Genes with fold change values greater than 1.5 (|log_2_ fold change| > 0.58) and a Benjamini-Hochberg adjusted *p* value ≤ 0.05 were considered DEGs. Analysis was performed on all IBM patients versus controls, male IBM patients versus male controls and female IBM patients versus female controls. Pathway enrichment analysis was carried out using the GSEA toolkit.^36, 37^ Spearman rank correlation analysis was performed for *NLRP3* against the whole dataset of IBM patients. To explore sex specific differences, we identified sex specific DEGs, defined as genes that were differentially expressed in one sex but not in the other, using the same cutoffs as described above. Genes with opposite trends were defined as those that were differentially expressed in both sexes but with opposite trends.

### Protein lysate preparation

For protein lysate preparation, cell lysis buffer from Cell Signaling (Danvers, MA, USA, cat. #9803) supplemented with PMSF, phosphatase and protease inhibitors, and 10-20 mg of fresh frozen muscle tissue were added to a tube with metal beads. Homogenization was performed using a Fisherbrand bead homogenizer followed by sonication. Centrifugation followed in a refrigerated (4°C) Beckman centrifuge. For protein quantification, we used the DC protein assay (Bio-Rad) following the manufacturer’s instructions.

### Western blot

A 6X sample buffer was added to the protein lysates followed by denaturation at 95°C. Total protein lysates (25 µg/well) were separated on 4–20% Criterion TGX™ Precast Protein Gels (Bio-Rad, 4561096) and transferred to a PVDF membrane. The following primary antibodies were used: NLRP3 (1:390, Novus Biologicals, NBP2-12446), ASC (1:500, Cell Signaling, E1E3I), vinculin (1:1000, Cell Signaling, Danvers, MA, USA, cat. #4650), and actin (1:1000, Cell Signaling, D18C11).

The following secondary antibodies were used: peroxidase (HRP), anti-rabbit IgG (H+L), goat secondary antibody (1:5000, Jackson ImmunoResearch, 111-035-003), and rabbit TrueBlot, anti-rabbit IgG HRP (1:5000, Rockland Immunochemicals, 18-8816-31). Membrane development was performed using a ChemiDoc Imaging System from Bio-Rad. Quantification was performed using ImageJ software.

### P-S65-Ub measurement via Meso Scale Discovery-based sandwich ELISA

Sandwich ELISA targeting p-S65-Ub was performed via the Meso Scale Discovery (MSD) platform that uses electrochemiluminescence (ECL), as described previously.^38, 39^ Briefly, 96-well plates (Meso Scale Diagnostics, L15XA-3) were coated overnight with 1 µg/ml capture p-S65-Ub antibody (Cell Signaling Technology, 62802) in 200 mM sodium carbonate buffer pH 9.7. MSD plates were washed twice with washing buffer (150 mM Tris, pH 7.4, 150 mM NaCl, 0.1% [v:v] Tween-20). Plates were then blocked with 1% BSA (w:v) in wash buffer for 1 hour. 30 µg of samples were prepared in blocking buffer and incubated for 2 h. Detergent volumes were kept consistent across all samples and the samples were run in duplicates.

Mouse pink1-/- skeletal muscle lysates were used as negative controls. Plates were washed three times and incubated with 5 µg/ml total Ub antibody (Thermo Fisher, 14-6078-37) for 2 h. Plates were washed and incubated with 1 µg/ml SULFO-TAG labeled goat anti-mouse antibody (MSD, R32AC-1) for 1 h. Plates were washed and measured immediately after addition of 150 μl MSD GOLD Read Buffer (MSD, R92TG-2) on a MESO QuickPlex SQ 120 reader (Meso Scale Diagnostics, Rockville, MD, USA).

### Muscle histopathology

10 µm frozen muscle sections were stained with hematoxylin and eosin, ATPase at pH 4.3 and 4.6, cytochrome c oxidase, NLRP3 (Novus Biologicals, NBP2-12446) and p-S65 Ub antibodies. The p-S65-Ub antibody was developed in house and fully characterized and validated in cells and human tissue.^40, 41^ Immunohistochemistry was performed on adjacent 10 μm sections of fresh-frozen human muscle fixed in cold acetone. p-S65-Ub stained sections were quenched in 0.3% hydrogen peroxide with methanol solution and rinsed with 1X PBS-X, and all sections were blocked with normal donkey serum (1:10, Jackson ImmunoResearch, 017-000-121). Primary antibodies were incubated overnight at 4°C, at concentrations of 1:650 p-S65-Ub and 1:50 NLRP3, followed by incubation with a biotinylated donkey anti-rabbit secondary antibody (1:500, Jackson ImmunoResearch, 711-065-152). Stains were visualized using Vectastain ABC (Vector Laboratories, PK-6100) and Dako DAB chromogen (Agilent, K346811-2). Slides were reviewed by E.N. (muscle pathologist) and photos were taken with an Olympus DP73 camera.

### Statistical analysis

Continuous data are represented as mean (SD) or median (interquartile range) as appropriate, and categorical variables are represented as proportions and percentages. Statistical analysis was performed using BlueSky statistics v.7.10 software, GraphPad Prism v.9.3.1 and SAS 9.4 software (SAS Inc, Cary NC) unless otherwise stated (refer to the transcriptomics section for transcriptomic analysis details). Two-tailed Mann-Whitney test and Pearson or Spearman correlation were used as appropriate.

### Data sharing statement

The Raw RNA sequencing data will be uploaded to a public database. Additional data supporting the findings of this study are available from the corresponding author upon reasonable request.

## Results

### Study cohort characteristics

The study population consisted of 38 IBM patients and 22 controls without myopathy, with equal proportion of males and females in each group **(Table 1)**. The mean age at biopsy was 62.9 years in the IBM group and 59.7 years in the control group. Mean disease duration at the time of biopsy in the IBM group was 6 years, and mean MMT score was 16.6.

**Table 1:** Baseline characteristics and patient demographics.

| Variable | Inclusion body myositis (n = 38) | Controls (n = 22) |
| --- | --- | --- |
| **Age at biopsy (years)** | 62.6 (9) | 59.7 (10) |
| **Sex:** |  |  |
| Females | 19 (50) | 11 (50) |
| Males | 19 (50) | 11 (50) |
| **Race:** |  |  |
| American Indian/Alaskan Native | 1 (2.6) | 1 (4.5) |
| Black | 1 (2.6) | 0 |
| White | 32 (84.2) | 19 (86.4) |
| Unknown/not disclosed | 4 (10.5) | 2 (9.1) |
| **Biopsy target** |  |  |
| Quadriceps | 14 (36.8) | 8 (36.4) |
| Biceps brachii | 9 (23.7) | 5 (22.7) |
| Triceps brachii | 10 (26.3) | 3 (13.6) |
| Others | 5 (13.2) | 6 (27.3) |
| **Disease characteristics** |  |  |
| Disease duration at time of the biopsy (years) | 6.1 (4) | NA |
| Manual Motor Test score | 16.6 (9.4) | 0 |
| Patients requiring any type of walking assistance at time of the biopsy | 4 (10.5) | 0 |

Continuous variables are displayed as mean (SD) and categorical variables as number of patients (percentage). NA: not applicable.

### Transcriptomic analysis of samples from IBM patients versus controls

To explore the molecular pathways involved in IBM pathogenesis, we first compared the IBM group to controls, including both males and females. Principal component analysis (PCA) revealed clear separation between the two groups **(Figure 1A)**. There were 7448 differentially expressed genes (DEGs) in IBM: 5777 upregulated and 1671 downregulated **(Figure 1A)**. A complete list of DEGs can be found in **Supplemental Table 1**. GSEA pathway analysis revealed that the NLRP3 inflammasome was the most upregulated pathway in IBM, followed by the interferon signaling pathway. Several of the top 10 downregulated pathways were related to mitochondria (the TCA cycle and respiratory electron transport, mitophagy and protein localization) **(Figure 1B)**. To evaluate sex differences, we compared female IBM patients to female controls and male IBM patients to male controls. In females, there were 4743 upregulated and 1544 downregulated genes in IBM. In males, there were 4993 upregulated and 1623 downregulated genes in IBM. A complete list of DEGs in males and females can be found in **Supplemental Tables 2-3**. We then identified sex-specific DEGs as shown in **Figure 1C**. There were 1754 female-specific and 2083 male-specific genes, and 2 genes with opposite trends between sexes. GSEA pathway analysis of sex-specific genes **(Figure 1D)** showed that the most upregulated female-specific pathways were related to the response to stress, defense response and regulation of the immune response. Whereas, the most upregulated male-specific pathways were related to cell adhesion, cell migration and the Fc receptors for IgG-dependent phagocytosis. The most downregulated sex-specific pathways related to RNA metabolism and translation, and electron transport chain and oxidative phosphorylation in females, and protein localization, ubiquitination and degradation, and regulation of DNA repair in males. Only two genes had opposite trends **(Figure 1C)**: *TPRG1* (Tumor Protein P63 Regulated 1) was upregulated in males and downregulated in females and *TSPAN5* (Tetraspanin 5) was upregulated in females and downregulated in males. Information about the role of *TPRG1* is sparse. *TPRG1* promoted inflammation and activation of NF-КB signaling in a murine model of cystitis.^42^ *TSPAN5* mediates signal transduction events that play a role in the regulation of cell development, migration and senescence.^43^

**Figure 1:**
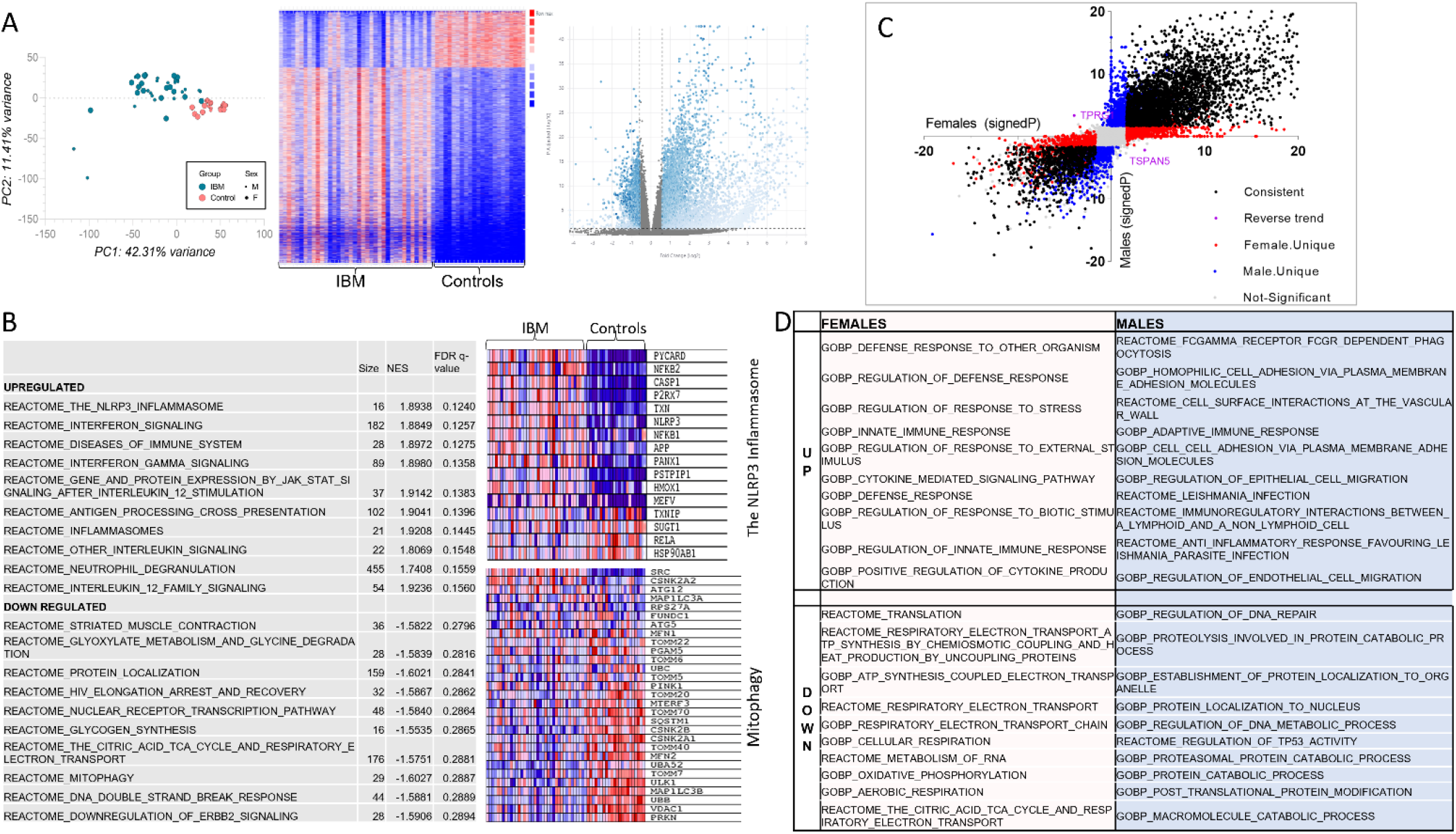
IBM transcriptome. Transcriptomic analysis of muscle tissue from 38 patients with inclusion body myositis and 22 controls. A) PCA (to the left) and heat map (middle) showing clear separation between IBM patients and controls. Volcano plot (to the right) representing differentially expressed genes (blue). B) Top 10 upregulated and top 10 downregulated pathways in IBM. The NLRP3 inflammasome and mitophagy pathways are detailed in the heatmaps with downregulated genes shown in blue and upregulated genes shown in red. Transcriptomic sex differences are shown in C&D. C) Volcano plot showing the signed -log_10_(*p* value) for each gene in females on the *x* axis and in males on *y* axis. Differentially expressed genes that are consistent in both sexes are shown in black, female-unique genes are shown in red, male-unique genes are shown in blue, and genes with reverse trends are shown in purple. Genes that did not reach fold change or *p*-value cutoffs in either sex are shown in gray (not significant). D) The top 10 upregulated and downregulated pathways for sex-specific genes.

### NLRP3 inflammasome activation in inclusion body myositis

Next, we validated the overall highest upregulated pathway, the NLRP3 inflammasome, at the protein level in muscle samples from 16 IBM patients and 10 controls by western blotting. There was increased expression of the two main NLRP3 inflammasome proteins: NLRP3 and ASC (apoptosis-associated speck-like protein containing CARD, also known as PYCARD: PYD and CARD Domain Containing) **(Figure 2A).** Upregulation of NLRP3-related genes and increased expression of NLRP3 and ASC proteins were observed in samples from both males and females. Immunohistochemistry demonstrated increased NLRP3 immunoreactivity in both inflammatory cells and scattered muscle fibers **(Figure 2B)**. Taken together, these data support that NLRP3 inflammasome is activated in IBM muscles consistent with the RNA-seq data.

**Figure 2:**
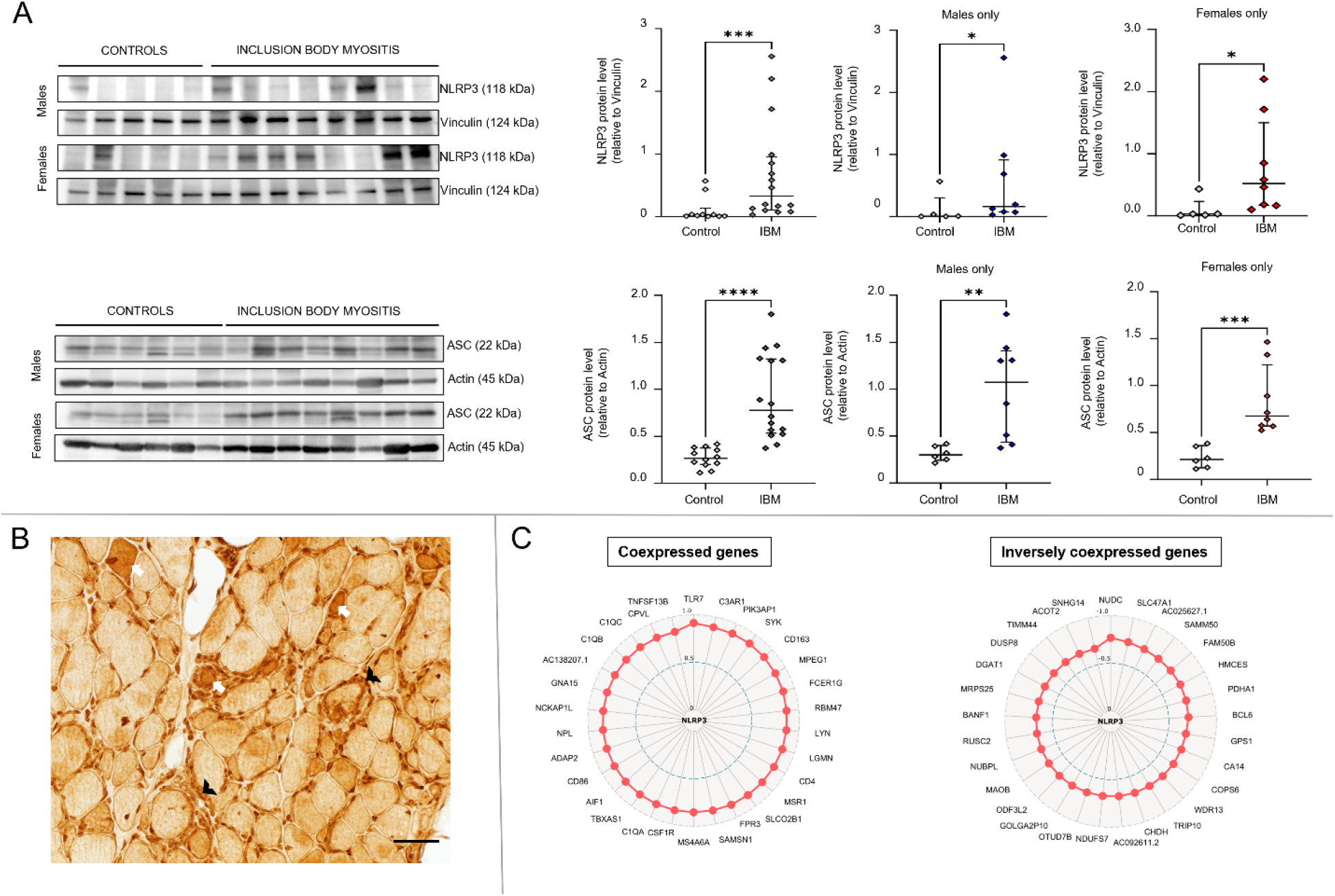
NLRP3 inflammasome activation in inclusion body myositis. A) Western blot of muscle lysates from IBM patients and controls showing increased NLRP3 and ASC protein expression in both males and females. Western blot quantification results comparing IBM group to controls were as follows (Mann-Whitney U, two-tailed *p*-value): NLRP3 for both sexes (20, *p =* 0.0009), NLRP3 for males (6, *p =* 0.045), NLRP3 for females (3, *p =* 0.011), ASC for both sexes (4, *p <* 0.0001), ASC for males (3, *p =* 0.0047), and ASC for females (0, *p =* 0.0007). B) NLRP3 immunohistochemistry showing increased NLRP3 immunoreactivity in scattered muscle fibers (examples shown as white arrows) as well as in inflammatory cells (examples shown as black arrowheads). C) Radar maps displaying the top coexpressed and inversely coexpressed genes with *NLRP3* using Spearman rank correlation analysis. The correlation coefficient corresponds to the distance from the center. All represented genes had a *p-value* less than 0.0001.

The aberrant activation and regulation of the NLRP3 inflammasome are intricate and there is a growing body of literature elucidating its complexities.^44^ To better understand the related genes in our dataset, coexpression analysis of *NLRP3* against all the other genes in the IBM dataset. The top 30 coexpressed and inversely coexpressed genes in IBM samples are shown in **Figure 2C**. Most of the coexpressed genes were related to the immune system/inflammation including *TLR7* (Toll Like Receptor 7) (highest ρ = 0.91), complement activation [*C3AR1* (Complement C3a Receptor 1)*, FPR3* (Formyl Peptide Receptor 3)*, C1QA* (Complement C1q A Chain)*, C1QB* (Complement C1q B Chain)*, C1QC* (Complement C1q C Chain)], as well as genes related to metabolism [*TBXAS1* (Thromboxane A Synthase 1)*, NPL* (N-Acetylneuraminate Pyruvate Lyase)], and calcium homeostasis [*GNA15* (G Protein Subunit Alpha 15)]. The top 30 inversely coexpressed genes included several mitochondria-associated genes related to the mitochondrial inner and outer membranes [*SAMM50* (SAMM50 Sorting And Assembly Machinery Component)*, MAOB* (Monoamine Oxidase B)*, TIMM44* (Translocase Of Inner Mitochondrial Membrane 44)], metabolism [*PDHA1* (Pyruvate Dehydrogenase E1 Subunit Alpha 1)*, CHDH* (Choline Dehydrogenase)*, ACOT2* (Acyl-CoA Thioesterase 2)], complex I [*NDUFS7* (NADH:Ubiquinone Oxidoreductase Core Subunit S7)], and mitochondrial protein synthesis [*MRPS25* (Mitochondrial Ribosomal Protein S25)]. In addition, there were genes related to protein ubiquitination and degradation [*COPS6* (COP9 Signalosome Subunit 6)*, SNHG14I* (Small Nucleolar RNA Host Gene 14)] and several either unidentified genes or genes of unknown functions. The gene with the strongest inverse correlation was *NUDC* (Nuclear Distribution C, Dynein Complex Regulator) (ρ = −0.77) related to cell division. *OTUD7B* (OTU Deubiquitinase 7B) is a regulator of noncanonical NF-κB activation.

### Altered mitophagy in muscle tissue from inclusion body myositis patients

Mitophagy was among the top 10 downregulated pathways in IBM in the combined male and female analysis, and mitophagy plays an important role in regulating inflammation in general and NLRP3 inflammasome activation in particular.^45^ To further explore whether mitophagy is altered in IBM muscles, we measured p-S65-Ub levels in muscle lysates from 22 IBM patients and 10 controls (**Figure 3A)**. The level of p-S65-Ub was significantly greater in the IBM group compared to controls (*p =* 0.005), indicating altered mitophagy. This can be due to either stronger mitophagy initiation or lower mitophagic degradation in IBM skeletal muscle. Analysis by sex revealed that compared with male controls, IBM males had significantly greater level of p-S65-Ub (*p* = 0.014), whereas females only showed mild non-significant increase compared to female controls (*p =* 0.31). To further explore the distribution of the findings at the tissue level, we performed muscle enzymatic and immunohistochemical studies **(Figure 3B-C)**. In muscle tissue from controls, type 2 fibers (fast twitch, lower mitochondrial content) had weak immunoreactivity to p-S65-Ub antibodies, while type 1 fibers (slow twitch, higher mitochondrial content) strongly reacted. This might reflect the difference in baseline mitophagy which is likely associated with the overall difference in mitochondrial content between fiber types. In contrast to samples from controls, there were p-S65-Ub positive aggregates in scattered muscle fibers in IBM muscles. These fibers were type 2 fibers that were also devoid of CCO activity (CCO negative fibers). Type 2 fibers are known to be more prone to mitochondrial failure.^46^ Also noted, all CCO negative fibers, with or without observable p-S65-Ub aggregates, were type 2 fibers.

**Figure 3:**
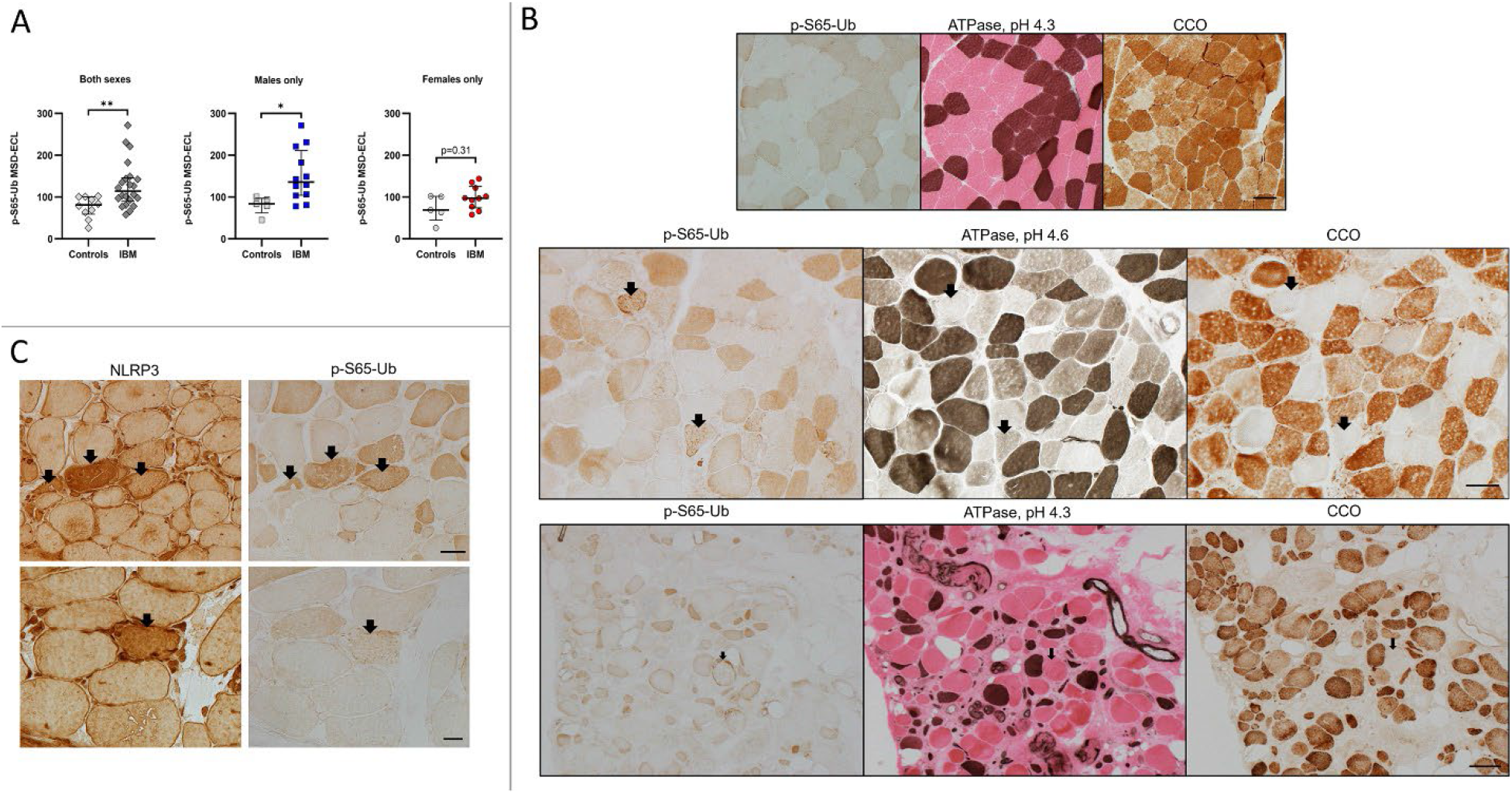
Altered mitophagy in inclusion body myositis patients. A) Measurement of p-S65-Ub levels via sandwich ELISA in muscle samples from IBM patients and controls. Results comparing IBM samples to controls were as follows (Mann-Whitney U, two-tailed *p*-value): both sexes (43, *p =* 0.0054), males only (7, *p =* 0.0136), and females only (16, *p =* 0.31). MSD-ECL: Meso Scale Discovery-Electrochemiluminescence. B&C) Frozen muscle sections from IBM samples and controls reacted to p-S65-Ub, NLRP3, ATPase at pH 4.3 (type 1 fibers: dark brown, type 2 fibers: pink) and at pH 4.6 (type 1 fibers: dark brown; type 2a fibers: light brown; type 2b fibers: in between shade), and cytochrome c oxidase. MSD: Mesoscale discovery; B) Top row: muscle samples from controls, p-S65-Ub shows clear differentiation of fiber types, reflecting the difference in baseline mitophagy and mitochondrial content between fiber types. Type 1 fibers, dark brown on ATPase 4.3, had increased p-S65-Ub and CCO reactivity. Middle and bottom rows: muscle samples from a male and a female IBM patients respectively, demonstrating p-S65-Ub positive aggregates in scattered muscle fibers (examples shown as arrows). These fibers were almost exclusively type 2 fibers (ATPase stains) that were also devoid of CCO activity (CCO negative fibers). Also noted, all CCO negative fibers, with or without observable abnormal p-S65-Ub reactivity, were type 2 fibers. Scale bar for panel B: top row: 50 µm; middle row: 50 µm; bottom row: 200 µm. C) Muscle samples from IBM patients, reacted for NLRP3 and p-S65-Ub, demonstrating muscle fibers with increased NLRP3 and p-S65-Ub reactivity (arrows). Scale bar: top row 50 µm, bottom row 20 µm

These data suggest that mitophagy alterations occur mostly in type 2 fibers in IBM and are more significant in males. Additionally, several fibers with increased NLRP3 signal also exhibited increased p-S65-Ub immunoreactivity **(Figure 3C)**, suggesting that these two pathways might be functionally associated.

### Correlation of NLRP3 inflammasome activation with altered mitophagy and muscle weakness

We next determined whether the *NLRP3* level or p-S65-Ub level was correlated with disease duration or disease severity, as reflected by the MMT score. The results are shown in **Figure 4**. The *NLRP3* level had a modest correlation with the p-S65-Ub level in both males and females (both sexes: correlation coefficient ρ = 0.41, males: ρ = 0.48, females: ρ = 0.54). *NLRP3* levels strongly correlated with MMT scores in males (ρ = 0.62) but not in females (ρ = −0.14). Neither the *NLRP3* nor the p-S65-Ub level correlated with disease duration. These findings further support the relationship between NLRP3 activation and altered mitophagy in IBM patients. Furthermore, NLRP3 activation is strongly associated with muscle weakness in males.

**Figure 4:**
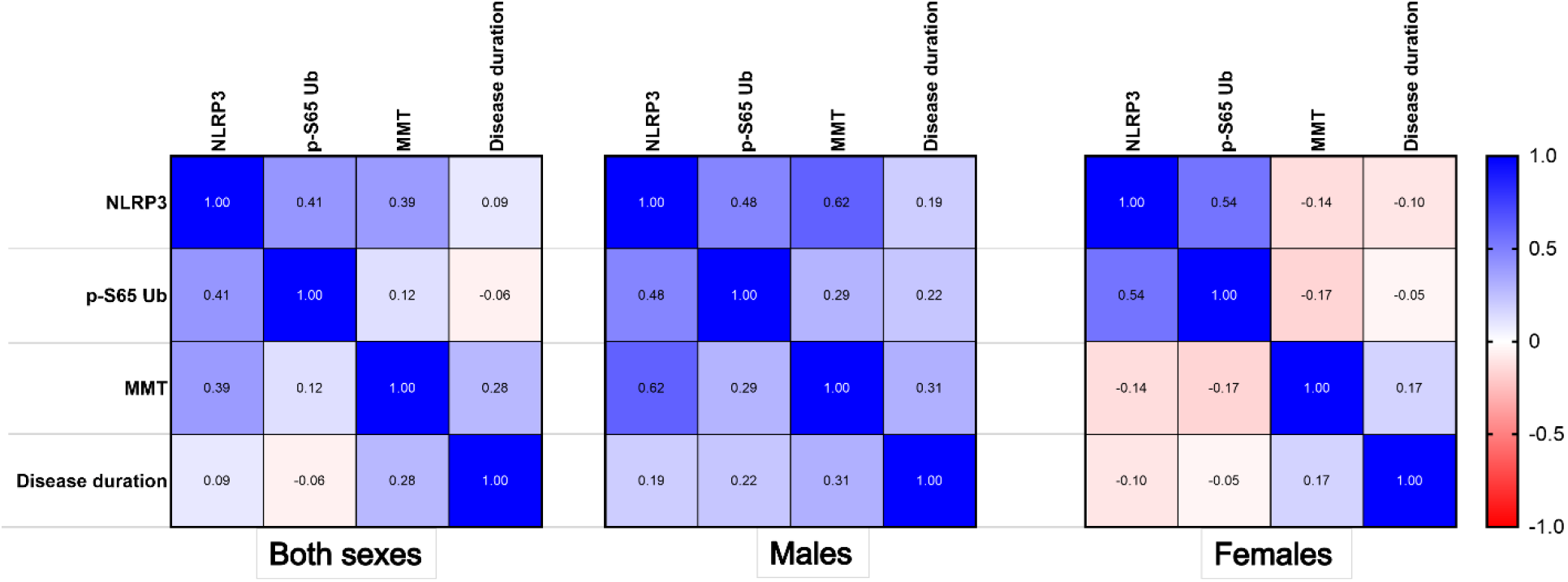
Correlation of NLRP3 inflammasome activation with altered mitophagy and muscle weakness. Pearson correlation plot demonstrating correlation coefficient values for the *NLRP3 RNA* level, p-S65-Ub level, manual muscle testing (MMT) score and disease duration, by sex. **Both sexes:** *NLRP3* and p-S65-Ub, *p =* 0.059; *NLRP3* and MMT score, *p =* 0.072; *NLRP3* and disease duration, *p =* 0.543; p-S65-Ub and MMT score, *p =* 0.58; p-S65-Ub and disease duration, *p =* 0.699. **Males:** *NLRP3* and p-S65-Ub, *p =* 0.116; *NLRP3* and MMT score, *p =* 0.032; *NLRP3* and disease duration, *p =* 0.699; p-S65-Ub and MMT score, *p =* 0.359; p-S65-Ub and disease duration, *p =* 0.49. **Females:** *NLRP3* and p-S65-Ub, *p =* 0.106; *NLRP3* and MMT score, *p =* 0.706; *NLRP3* and disease duration, *p*-=0.781); p-S65-Ub and MMT score, *p =* 0.642; p-S65-Ub and disease duration, *p =* 0.898.

## Discussion

Our study, for the first time, provides preliminary evidence that a vicious cycle of NLRP3 inflammasome activation and altered mitophagy exists in IBM muscles. Despite their intended protective role, aberrant activation of inflammasomes has been linked to the development of various chronic conditions, especially neurodegenerative diseases and other diseases of aging.^44, 47^ The inflammasomes are supramolecular platforms for the activation of caspases through the release of proinflammatory cytokines IL-1β and IL-18, resulting in programmed cell death or apoptosis.^48^ NLRP3 remains the most studied inflammasome and has been implicated in the pathogenesis of many neurodegenerative diseases of aging such as AD, PD, and amyotrophic lateral sclerosis, but studies on its role in muscle disorders are limited.^44, 47, 49^ In a recent study on IBM, Kummer et al. demonstrated increased *NLRP3* mRNA levels by targeted qt-PCR in muscle tissue from IBM patients and postulated that NLRP3 may be a key driver of inflammation and protein accumulation in IBM.^50^ Herein, we demonstrated via unbiased analysis of transcriptomic data that the NLPR3 inflammasome pathway was indeed the most upregulated pathway in muscle tissue from both males and females with IBM with increased NLRP3 and ASC protein expression. The NLRP3 inflammasome belongs to the NOD-like receptor (NLR) signaling pathway which was the most significantly upregulated pathway in IBM muscle in a study by Murakami et al., validating our results.^27^ However, the authors did not provide further details on the mechanism of NLR signaling pathway and it was grouped under “infection pathways”. Similarly, the “neuroinflammation signaling pathway”, which includes the NLRP3 inflammasome, was the most significantly upregulated pathway in IBM in a study by Pinal Fernandez et al.^24^ It is noteworthy that correlation between histopathological and clinical characteristics is typically weak in IBM, given complexity of the disease and the heterogeneity of the IBM population.^33^ However, we demonstrate strong correlation between the *NLRP3* RNA level and the severity of muscle weakness, reflected by the MMT score, in males but not in females, supporting the clinical relevance of the activation of the NLRP3 inflammasome in IBM.

Mitophagy serves to remove dysfunctional mitochondria via the autophagic machinery to maintain cellular homeostasis and mitochondrial integrity. While the PINK1-PRKN mitophagy pathway is genetically linked to PD, both proteins are expressed throughout the body and this pathway likely plays an important role beyond the brain. However, there are limited studies on mitophagy in human skeletal muscles. PRKN is thought to play an important role in the maintenance of mitochondrial integrity in skeletal muscles, and aging is associated with a decline in mitophagy, mitochondrial function and muscle mass.^51^ Herein, for the first time, we show robust alterations in the PINK1-PRKN mitophagy pathway in IBM. Elevated p-S65-Ub levels could stem from the accumulation of damaged mitochondria or from a block in the degradation of labeled mitochondria. Interestingly, the changes in p-S65-Ub were pronounced in males and more research is needed to determine sex-specific changes in PINK1-PRKN mitophagy mechanism(s). At the tissue level, altered mitophagy was observed in CCO-negative fibers. We also showed that loss of CCO activity and altered mitophagy both occur mainly in type 2 fibers. Type 2 fibers are more prone to mitochondrial failure because they have lower mitochondrial content and lower oxidative capacity being fast-twitch fibers that rely on glycolysis, and exhibit lower expression of E3 Ub-ligases and proteasome-mediated protein degradation.^46^ CCO negative fibers in IBM have also been shown to have increased mitochondrial DNA mutation loads.^16^ Moreover, the predilection of type 2 fibers also raises suspicion for underlying metabolic disturbances, which has not yet been fully explored in IBM. Lastly, unbiased coexpression analysis demonstrating that genes with strongest inverse correlation with *NLRP3* level were mostly related to the mitochondria, immunohistochemical studies demonstrating abnormal p-S65-Ub aggregates in fibers with increased NLRP3 immunoreactivity, and the correlation between the p-S65-Ub level and the *NLRP3* level support that these pathways are functionally related in IBM. Based on these results, a vicious cycle of mitochondrial dysfunction/altered mitophagy and NLRP3 inflammasome activation is likely to occur in IBM (**Figure 5)**. The release of mitochondrial damage-associated molecular patterns (DAMPs) such as mtDNA, cytochrome c, mitochondrial reactive oxygen species (ROS), and mitochondria-specific cardiolipin molecules, are known activators of the NLRP3 inflammasome.^44^ Under normal conditions, damaged mitochondria and the NRLP3 inflammasome are both removed by mitophagy and autophagy subsequently, reestablishing cellular homeostasis.^45, 52^ When mitophagy and autophagy are altered, as in IBM, a feedforward loop is established.^45, 53^ Thereby, the inflammatory milieu results in additional oxidative stress and mitochondrial dysfunction with further release of mitochondrial DAMPs and subsequent aberrant NLRP3 inflammasome activation. This vicious self-sustaining cycle may be a major contributor to the chronic deterioration of muscle strength in individuals with IBM. Although not directly assessed in our study, the 26S proteasome and autophagy, the two major pathways for protein degradation in eukaryotic cells, have both been shown to be impaired in IBM.^54^ Additionally, decreased lysosomal proteolytic activity despite increased maturation of autophagosomes, as well as the upregulation and aggregation of chaperone-mediated autophagy components have been demonstrated in IBM muscles.^55, 56^ The accumulation of autophagic vacuoles and p62+ aggregates in muscle samples is used to aid in the histopathological diagnosis of IBM; p62 is a shuttle protein that transports polyubiquitinated proteins to either proteasomal or lysosomal degradation.^32, 57^ The original trigger for this self-sustained detrimental loop remains unclear. An aging skeletal milieu, with aging mitochondria and autophagic system, is probably required for these events to occur given the age of the population at risk.^13^ Furthermore, antigen-driven inflammation at a preclinical stage preceding the activation of self-sustained inflammatory pathways that become nontargeted by conventional immunotherapies is possible. Environmental and genetic risk factors could also be potential contributors to IBM pathogenesis.^58^

**Figure 5:**
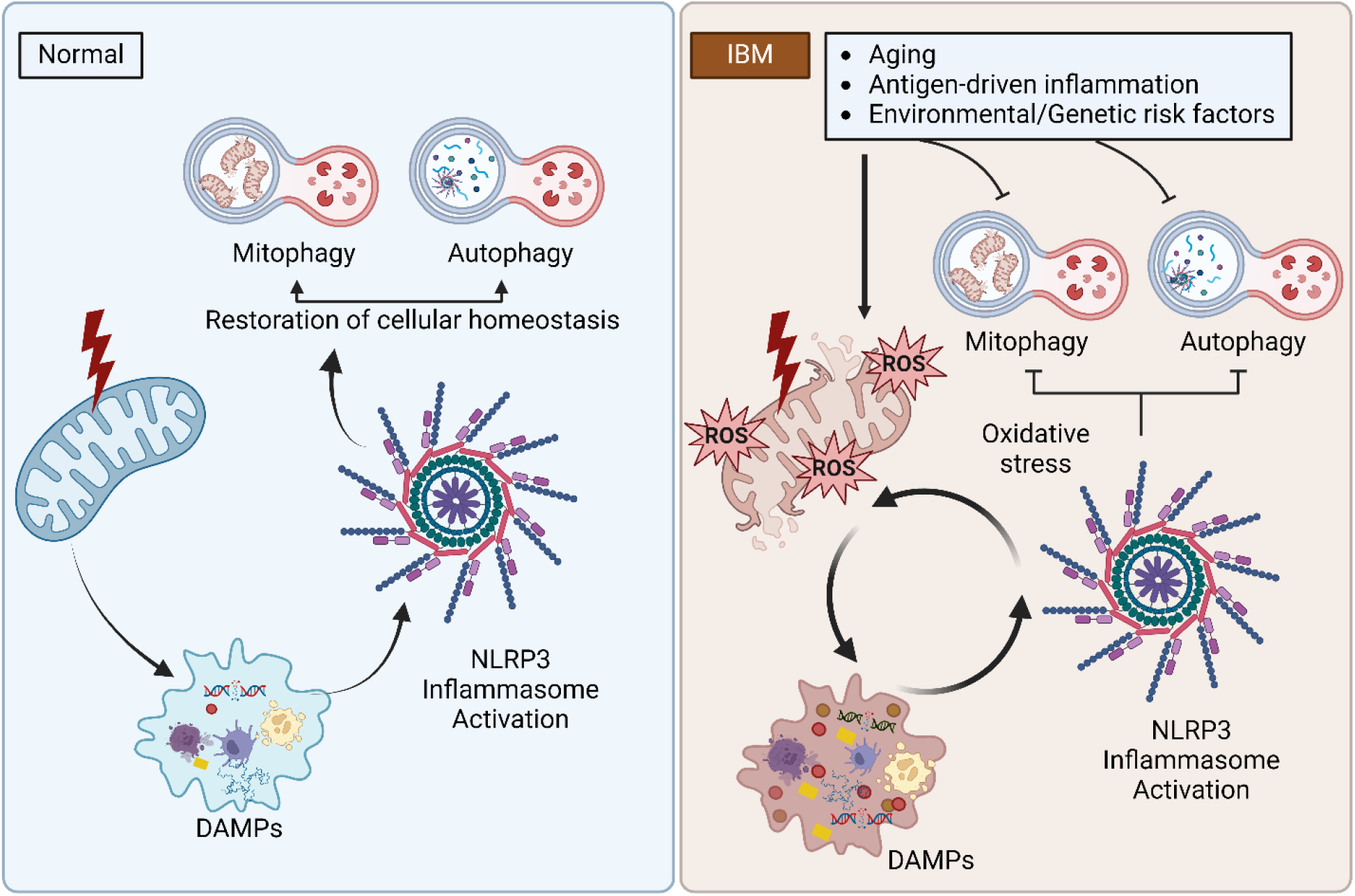
The vicious cycle of inflammasome activation-mitochondrial dysfunction/altered mitophagy in inclusion body myositis. The release of mitochondrial damage-associated molecular patterns (DAMPs) results in the activation of the NLRP3 inflammasome. Under normal conditions, damaged mitochondria and the NRLP3 inflammasome are both subsequently removed by mitophagy and autophagy, reestablishing cellular homeostasis. In IBM, mitophagy and autophagy are altered, establishing a feedforward loop in which the inflammatory milieu results in additional oxidative stress and mitochondrial dysfunction with further release of mitochondrial DAMPs and subsequent aberrant NLRP3 inflammasome activation.

Another major finding of this study is the demonstration of sex differences in IBM pathomechanisms. Male sex is a known risk factor for IBM as males are twice as likely to have the disease than females.^29, 31^ Regarding disease severity, males are reported to have more significant limb weakness.^30^ Furthermore, additional variations in phenotype by sex have been described.^34^ Despite these phenotypic sex differences, none of the previous studies explored underlying differential disease mechanisms. In this study, we identified sex-specific genes and pathways that were differentially expressed only in one sex. Interestingly, the upregulated pathways in females could conceivably be protective likely indicating that females fared to respond better to stress and to the immune system activation, whereas in males, the upregulated pathways of cell adhesion and migration might have promoted disease pathogenesis. Increased expression of cell adhesion molecules, such as cadherin 1 has been reported in IBM.^28^ For downregulated sex-specific genes and related pathways, it is possible that females have decreased oxidative phosphorylation as a protection from oxidative stress, or as a result of lower mitochondrial mass from more preserved mitophagy. In contrast, pathways related to protein homeostasis and localization were downregulated in males. Disrupted protein homeostasis with the aggregation of autophagic vacuoles and misfolded protein aggregates is a canonical histopathological feature of IBM.^59, 60^ Last, males had more significantly altered mitophagy than females, and *NLRP3* levels correlated with the severity of muscle weakness in males supporting a direct association between NLPR3 activation and loss of muscle strength in this disease. Taken together, these observations shed light on potential mechanistic differences between sexes, possibly explaining, at least in part, why males are more commonly and severely affected in IBM, and offer solid hypotheses to be explored in future studies.

Limitations of this study include the retrospective nature of the chart review, and the lack of a valid disease model for IBM to further investigate the underlying pathomechanisms.^9^ The main strength of the study is that the experiments were performed on muscle samples from IBM patients providing insight into its pathogenesis. However, there is some heterogeneity inherent to the nature of the disease, stage of the disease at diagnosis, and muscle biopsy target.

The findings of this study may have major therapeutic implications. Small molecule mitochondria-targeted therapeutics are being investigated for the treatment of neurodegenerative diseases of aging.^61^ Intervening on mitochondrial pathways requires characterizing the specific level of mitochondrial dysfunction in a particular disorder.^62, 63^ Furthermore, there has been rapid advancement in developing strategies for inflammasome blockade, and targeting both mitochondrial dysfunction and inflammasome activation in neurodegenerative diseases such as PD and AD.^64–66^ Deciphering the intertwined nature of neurodegenerative diseases resulting in self-sustaining vicious cycles, such as those involving inflammasome activation and stalled mitophagy, has become of utmost importance, regardless of which is the cause versus the consequence. How to simultaneously target the various interwoven pathways involved in neurodegenerative diseases remains unknown and should be investigated in future studies.

## ACKNOWLEDGEMENTS

We thank Surendra Dasari for his technical help in creating the radar chart for coexpression analysis. This research was supported by grants from the National Institute of Arthritis, Musculoskeletal and Skins Diseases (NIAMS, K08-AR78254), American Neuromuscular Foundation (ANF #2, career development grant), and Mayo Clinic Center for Clinical and Translational Sciences (small project grant and Team Science pilot award, UL1TR002377 from the National Center for Advancing Translational Sciences) to E.N. W.S. is partially supported by grants from the National Institute of Neurological Disorders and Stroke (RF1 NS085070, R01 NS110085, and U54 NS110435), the Michael J. Fox Foundation for Parkinson’s Research, the Ted Nash Long Life Foundation, Mayo Clinic Foundation, Mayo Clinic Center for Biomedical Discovery, and Mayo Clinic Robert and Arlene Kogod Center on Aging. F.C.F. is supported by grants from the Florida Department of Health (22A07), the Michael J Fox Foundation for Parkinson’s Research, and the Mayo Clinic Center for Biomedical Discovery. X.H. is supported by a pilot grant and a developmental project award from the Mayo Clinic Alzheimer Disease Research Center (ADRC, P30 AG062677) and fellowships awarded by the APDA and Alzheimer’s Association [AARF-22-973152]. E.T., H.G. and T.K.O.N. were supported by a grant from the National Institutes of Health (RF1AG55549 to E.T.). The study contents are solely the responsibility of the authors and do not necessarily represent the official view of the NIH and other funding organizations. The funders had no role in the study design, data collection and analysis, the decision to publish, or the preparation of the manuscript.

## Author contributions

Conceptualization of the study: E.N.; study design: E.N., W.S., E.T.; RNA sequencing data analysis: E.N., H.G., J.M.; co-expression analysis: W.S.H., J.M.; performance of meso-scale-discovery ELISA: J.O.W., conduction of Western Blot: E.N., T.K.O.N., immunohistochemistry: E.N., E.K.; writing of the first manuscript draft: E.N.; data interpretation: E.N., T.K.O.N., J.O.W., X.H., F.C.F., I.L., W.S., E.T.; revision of the manuscript for intellectual content: all authors; study supervision: E.N., W.S., E.T.. All authors have agreed to the published version of the manuscript.

## Declarations

## Ethical statement

The study was approved by Mayo Clinic Institutional Review Board. The study was considered minimal risk; therefore, the requirement for informed consent was waived. However, records of any patient who had not provided authorization for their medical records to be used for research, as per Minnesota statute 144.335, were not reviewed. The use of fresh-frozen clinical residual muscle biopsy tissue was also approved by the Mayo Clinic Biospecimens Subcommittee.

## REFERENCES

1. Weihl CC. Sporadic Inclusion Body Myositis and Other Rimmed Vacuolar Myopathies. Continuum (Minneapolis, Minn) 2019;25:1586–1598.

2. Naddaf E. Inclusion body myositis: Update on the diagnostic and therapeutic landscape. Front Neurol 2022;13:1020113.

3. Shelly S, Mielke MM, Mandrekar J, et al. Epidemiology and Natural History of Inclusion Body Myositis: A 40-Year Population-Based Study. Neurology 2021.

4. Naddaf E, Shelly S, Mandrekar J, et al. Survival and associated comorbidities in inclusion body myositis. Rheumatology (Oxford, England) 2022;61:2016–2024.

5. Engel AG, Arahata K. Monoclonal antibody analysis of mononuclear cells in myopathies. II: Phenotypes of autoinvasive cells in polymyositis and inclusion body myositis. Ann Neurol 1984;16:209–215.

6. Amemiya K, Granger RP, Dalakas MC. Clonal restriction of T-cell receptor expression by infiltrating lymphocytes in inclusion body myositis persists over time. Studies in repeated muscle biopsies. Brain 2000;123 ( Pt 10):2030–2039.

7. Greenberg SA, Pinkus JL, Kong SW, Baecher-Allan C, Amato AA, Dorfman DM. Highly differentiated cytotoxic T cells in inclusion body myositis. Brain 2019;142:2590–2604.

8. Callan A, Capkun G, Vasanthaprasad V, Freitas R, Needham M. A Systematic Review and Meta-Analysis of-Prevalence Studies of Sporadic Inclusion Body Myositis. Journal of neuromuscular diseases 2017;4:127–137.

9. Skolka MP, Naddaf E. Exploring challenges in the management and treatment of inclusion body myositis. Current opinion in rheumatology 2023;35:404–413.

10. Lindgren U, Pullerits R, Lindberg C, Oldfors A. Epidemiology, Survival, and Clinical Characteristics of Inclusion Body Myositis. Ann Neurol 2022;92:201–212.

11. Salajegheh M, Pinkus JL, Taylor JP, et al. Sarcoplasmic redistribution of nuclear TDP-43 in inclusion body myositis. Muscle Nerve 2009;40:19–31.

12. Mendell JR, Sahenk Z, Gales T, Paul L. Amyloid filaments in inclusion body myositis. Novel findings provide insight into nature of filaments. Archives of neurology 1991;48:1229–1234.

13. Askanas V, Engel WK, Nogalska A. Sporadic inclusion-body myositis: A degenerative muscle disease associated with aging, impaired muscle protein homeostasis and abnormal mitophagy. Biochim Biophys Acta 2015;1852:633–643.

14. Chahin N, Engel AG. Correlation of muscle biopsy, clinical course, and outcome in PM and sporadic IBM. Neurology 2008;70:418–424.

15. Dahlbom K, Lindberg C, Oldfors A. Inclusion body myositis: morphological clues to correct diagnosis. Neuromuscul Disord 2002;12:853–857.

16. Oldfors A, Moslemi AR, Fyhr IM, Holme E, Larsson NG, Lindberg C. Mitochondrial DNA deletions in muscle fibers in inclusion body myositis. Journal of neuropathology and experimental neurology 1995;54:581–587.

17. Hedberg-Oldfors C, Lindgren U, Basu S, et al. Mitochondrial DNA variants in inclusion body myositis characterized by deep sequencing. Brain pathology (Zurich, Switzerland) 2021;31:e12931.

18. Rygiel KA, Miller J, Grady JP, Rocha MC, Taylor RW, Turnbull DM. Mitochondrial and inflammatory changes in sporadic inclusion body myositis. Neuropathol Appl Neurobiol 2015;41:288–303.

19. Truban D, Hou X, Caulfield TR, Fiesel FC, Springer W. PINK1, Parkin, and Mitochondrial Quality Control: What can we Learn about Parkinson’s Disease Pathobiology? J Parkinsons Dis 2017;7:13–29.

20. Chatzinikita E, Maridaki M, Palikaras K, Koutsilieris M, Philippou A. The Role of Mitophagy in Skeletal Muscle Damage and Regeneration. Cells 2023;12.

21. Ordureau A, Sarraf SA, Duda DM, et al. Quantitative proteomics reveal a feedforward mechanism for mitochondrial PARKIN translocation and ubiquitin chain synthesis. Mol Cell 2014;56:360–375.

22. Cortese A, Plagnol V, Brady S, et al. Widespread RNA metabolism impairment in sporadic inclusion body myositis TDP43-proteinopathy. Neurobiology of aging 2014;35:1491–1498.

23. Pinal-Fernandez I, Casal-Dominguez M, Derfoul A, et al. Identification of distinctive interferon gene signatures in different types of myositis. Neurology 2019;93:e1193–e1204.

24. Pinal-Fernandez I, Casal-Dominguez M, Derfoul A, et al. Machine learning algorithms reveal unique gene expression profiles in muscle biopsies from patients with different types of myositis. Annals of the rheumatic diseases 2020;79:1234–1242.

25. Johari M, Vihola A, Palmio J, et al. Comprehensive transcriptomic analysis shows disturbed calcium homeostasis and deregulation of T lymphocyte apoptosis in inclusion body myositis. J Neurol 2022.

26. Parkes JE, Thoma A, Lightfoot AP, Day PJ, Chinoy H, Lamb JA. MicroRNA and mRNA profiling in the idiopathic inflammatory myopathies. BMC Rheumatology 2020;4:25.

27. Murakami A, Noda S, Kazuta T, et al. Metabolome and transcriptome analysis on muscle of sporadic inclusion body myositis. Ann Clin Transl Neurol 2022;9:1602–1615.

28. Ikenaga C, Date H, Kanagawa M, et al. Muscle Transcriptomics Shows Overexpression of Cadherin 1 in Inclusion Body Myositis. Ann Neurol 2022;91:317–328.

29. Dobloug GC, Antal EA, Sveberg L, et al. High prevalence of inclusion body myositis in Norway; a population-based clinical epidemiology study. European journal of neurology 2015;22:672–e641.

30. Michelle EH, Pinal-Fernandez I, Casal-Dominguez M, et al. Clinical Subgroups and Factors Associated With Progression in Patients With Inclusion Body Myositis. Neurology 2023.

31. Cox FM, Titulaer MJ, Sont JK, Wintzen AR, Verschuuren JJ, Badrising UA. A 12-year follow-up in sporadic inclusion body myositis: an end stage with major disabilities. Brain 2011;134:3167–3175.

32. Rose MR. 188th ENMC International Workshop: Inclusion Body Myositis, 2-4 December 2011, Naarden, The Netherlands. Neuromuscul Disord 2013;23:1044–1055.

33. Pinto MV, Laughlin RS, Klein CJ, Mandrekar J, Naddaf E. Inclusion body myositis: correlation of clinical outcomes with histopathology, electromyography and laboratory findings. Rheumatology (Oxford, England) 2022;61:2504–2511.

34. Alamr M, Pinto MV, Naddaf E. Atypical presentations of inclusion body myositis: Clinical characteristics and long-term outcomes. Muscle Nerve 2022;66:686–693.

35. Dobin A, Davis CA, Schlesinger F, et al. STAR: ultrafast universal RNA-seq aligner. Bioinformatics (Oxford, England) 2013;29:15–21.

36. Subramanian A, Tamayo P, Mootha VK, et al. Gene set enrichment analysis: a knowledge-based approach for interpreting genome-wide expression profiles. Proc Natl Acad Sci U S A 2005;102:15545–15550.

37. Mootha VK, Lindgren CM, Eriksson KF, et al. PGC-1alpha-responsive genes involved in oxidative phosphorylation are coordinately downregulated in human diabetes. Nat Genet 2003;34:267–273.

38. Watzlawik JO, Hou X, Fricova D, et al. Sensitive ELISA-based detection method for the mitophagy marker p-S65-Ub in human cells, autopsy brain, and blood samples. Autophagy 2021;17:2613–2628.

39. Watzlawik JO, Fiesel FC, Fiorino G, et al. Basal activity of PINK1 and PRKN in cell models and rodent brain. Autophagy 2023:1–12.

40. Fiesel FC, Ando M, Hudec R, et al. (Patho-)physiological relevance of PINK1-dependent ubiquitin phosphorylation. EMBO Rep 2015;16:1114–1130.

41. Hou X, Fiesel FC, Truban D, et al. Age- and disease-dependent increase of the mitophagy marker phospho-ubiquitin in normal aging and Lewy body disease. Autophagy 2018;14:1404–1418.

42. Hong T, Piao S, Sun L, Tao Y, Ke M. Tumor protein P63 Regulated 1 contributes to inflammation and cell proliferation of cystitis glandularis through regulating the NF-кB/cyclooxygenase-2/prostaglandin E2 axis. Bosn J Basic Med Sci 2022;22:100–109.

43. Schreyer L, Mittermeier C, Franz MJ, et al. Tetraspanin 5 (TSPAN5), a Novel Gatekeeper of the Tumor Suppressor DLC1 and Myocardin-Related Transcription Factors (MRTFs), Controls HCC Growth and Senescence. Cancers (Basel) 2021;13.

44. Litwiniuk A, Baranowska-Bik A, Domańska A, Kalisz M, Bik W. Contribution of Mitochondrial Dysfunction Combined with NLRP3 Inflammasome Activation in Selected Neurodegenerative Diseases. Pharmaceuticals (Basel) 2021;14.

45. Marchi S, Guilbaud E, Tait SWG, Yamazaki T, Galluzzi L. Mitochondrial control of inflammation. Nat Rev Immunol 2023;23:159–173.

46. van Wessel T, de Haan A, van der Laarse WJ, Jaspers RT. The muscle fiber type–fiber size paradox: hypertrophy or oxidative metabolism? European Journal of Applied Physiology 2010;110:665–694.

47. Péladeau C, Sandhu JK. Aberrant NLRP3 Inflammasome Activation Ignites the Fire of Inflammation in Neuromuscular Diseases. Int J Mol Sci 2021;22.

48. Bulté D, Rigamonti C, Romano A, Mortellaro A. Inflammasomes: Mechanisms of Action and Involvement in Human Diseases. Cells 2023;12.

49. Liu D, Xiao Y, Zhou B, et al. PKM2-dependent glycolysis promotes skeletal muscle cell pyroptosis by activating the NLRP3 inflammasome in dermatomyositis/polymyositis. Rheumatology (Oxford, England) 2021;60:2177–2189.

50. Kummer K, Bertram I, Zechel S, Hoffmann DB, Schmidt J. Inflammasome in Skeletal Muscle: NLRP3 Is an Inflammatory Cell Stress Component in Inclusion Body Myositis. Int J Mol Sci 2023;24.

51. Leduc-Gaudet JP, Picard M, St-Jean Pelletier F, et al. Mitochondrial morphology is altered in atrophied skeletal muscle of aged mice. Oncotarget 2015;6:17923–17937.

52. Wu M, Li H, He J, Liang J, Liu Y, Zhang W. TRIM72 Alleviates Muscle Inflammation in mdx Mice via Promoting Mitophagy-Mediated NLRP3 Inflammasome Inactivation. Oxidative medicine and cellular longevity 2023;2023:8408574.

53. Nakahira K, Haspel JA, Rathinam VA, et al. Autophagy proteins regulate innate immune responses by inhibiting the release of mitochondrial DNA mediated by the NALP3 inflammasome. Nature immunology 2011;12:222–230.

54. Fratta P, Engel WK, McFerrin J, Davies KJ, Lin SW, Askanas V. Proteasome inhibition and aggresome formation in sporadic inclusion-body myositis and in amyloid-beta precursor protein-overexpressing cultured human muscle fibers. The American journal of pathology 2005;167:517–526.

55. Nogalska A, D’Agostino C, Terracciano C, Engel WK, Askanas V. Impaired autophagy in sporadic inclusion-body myositis and in endoplasmic reticulum stress-provoked cultured human muscle fibers. The American journal of pathology 2010;177:1377–1387.

56. Cacciottolo M, Nogalska A, D’Agostino C, Engel WK, Askanas V. Chaperone-mediated autophagy components are upregulated in sporadic inclusion-body myositis muscle fibres. Neuropathol Appl Neurobiol 2013;39:750–761.

57. Nogalska A, Terracciano C, D’Agostino C, King Engel W, Askanas V. p62/SQSTM1 is overexpressed and prominently accumulated in inclusions of sporadic inclusion-body myositis muscle fibers, and can help differentiating it from polymyositis and dermatomyositis. Acta Neuropathologica 2009;118:407–413.

58. Miller FW, Lamb JA, Schmidt J, Nagaraju K. Risk factors and disease mechanisms in myositis. Nature reviews Rheumatology 2018;14:255–268.

59. Dubourg O, Wanschitz J, Maisonobe T, et al. Diagnostic value of markers of muscle degeneration in sporadic inclusion body myositis. Acta Myol 2011;30:103–108.

60. Ahmed M, Machado PM, Miller A, et al. Targeting protein homeostasis in sporadic inclusion body myositis. Sci Transl Med 2016;8:331ra341.

61. Cunnane SC, Trushina E, Morland C, et al. Brain energy rescue: an emerging therapeutic concept for neurodegenerative disorders of ageing. Nature reviews Drug discovery 2020;19:609–633.

62. Murphy MP, Hartley RC. Mitochondria as a therapeutic target for common pathologies. Nature reviews Drug discovery 2018;17:865–886.

63. Zheng C, Nguyen KK, Vishnivetskiy SA, Gurevich VV, Gurevich EV. Arrestin-3 binds parkin and enhances parkin-dependent mitophagy. J Neurochem 2024.

64. Vande Walle L, Lamkanfi M. Drugging the NLRP3 inflammasome: from signalling mechanisms to therapeutic targets. Nature Reviews Drug Discovery 2024;23:43–66.

65. Khot M, Sood A, Tryphena KP, et al. NLRP3 inflammasomes: A potential target to improve mitochondrial biogenesis in Parkinson’s disease. Eur J Pharmacol 2022;934:175300.

66. Wilkins HM, Swerdlow RH. Relationships Between Mitochondria and Neuroinflammation: Implications for Alzheimer’s Disease. Curr Top Med Chem 2016;16:849–857.

